# Vascular Comorbidities Worsen Prognosis of Patients with Heart Failure Hospitalized with COVID-19

**DOI:** 10.1101/2021.03.23.21254209

**Authors:** Jacob Mok, Juan Malpartida, Kimberly O’Dell, Joshua Davis, Cuilan Gao, Harish Manyam

## Abstract

**Background:** Prior diagnosis of heart failure (HF) is associated with increased length of hospital stay (LOS) and mortality from Coronavirus disease-2019 (COVID-19). Associations between substance use, venous thromboembolism (VTE), or peripheral arterial disease (PAD) and its effects on LOS or mortality in patients with HF hospitalized with COVID-19 remains unknown.

**Objective:** This study identified risk factors associated with poor in-hospital outcomes among patients with HF hospitalized with COVID-19.

**Methods:** Case control study was conducted of patients with prior diagnosis of HF hospitalized with COVID-19 at an academic tertiary care center from January 1, 2020 to February 28, 2021. Patients with HF hospitalized with COVID-19 with risk factors were compared with those without risk factors for clinical characteristics, length of stay (LOS), and mortality. Multivariate regression was conducted to identify multiple predictors of increased LOS and in-hospital mortality in patients with HF hospitalized with COVID-19.

**Results:** Total of 211 HF patients were hospitalized with COVID-19. Females had longer LOS than males (9 days vs. 7 days; p < 0.001). Compared with patients without peripheral arterial disease (PAD) or ischemic stroke, patients with PAD or ischemic stroke had longer LOS (7 days vs. 9 days; p = 0.012 and 7 days vs. 11 days, p < 0.001; respectively). Older patients (aged 65 and above) had increased in-hospital mortality compared to younger patients (Adjusted OR: 1.04; 95% CI: 1.00 – 1.07; p = 0.036). VTE increased mortality more than three-fold in patients with HF hospitalized with COVID-19 (Adjusted OR: 3.33; 95% CI: 1.29 – 8.43; p = 0.011).

**Conclusion:** Vascular diseases increase LOS and mortality in patients with HF hospitalized with COVID-19.

**KEY QUESTIONS:** *What is already known about this subject?:*

- Prior diagnosis of heart failure (HF) increases LOS and mortality in patients admitted to the hospital for COVID-19
- Antiplatelet, anticoagulation, and statin therapy decreased venous thromboembolism (VTE) in patients admitted for COVID-19

*What does this study add?:*

- This study showed that patients with COVID-19, HF, and VTE had a higher mortality rate than patients with COVID-19 and either HF or VTE, or patients with HF and/or VTE who did not have COVID-19.
- This study showed that patients with HF hospitalized with COVID-19 had greater length of stay with prior diagnosis of peripheral arterial disease (PAD) or ischemic stroke

*How might this impact on clinical practice?:*

- Our findings demonstrate clinical relevance by showing supportive evidence for antiplatelet, anticoagulation, and statin therapy in HF patients hospitalized with COVID-19

## INTRODUCTION

The coronavirus disease-2019 (COVID-19) remains a deadly pandemic that is causing significant morbidity and mortality throughout the world. In spite of this, heart failure (HF) remains one of the most common diagnoses for hospital inpatient stays during the pandemic. A recent study showed that hospital LOS and risk of mortality were increased for patients hospitalized with COVID-19 with a prior diagnosis of HF, regardless of heart failure type,[1]. However, there is currently limited data on how different types of comorbidities impact the LOS and mortality of HF patients hospitalized with COVID-19. Furthermore, the effect of substance use, peripheral arterial disease (PAD), and venous thromboembolism (VTE) on LOS and mortality in HF patients hospitalized with COVID-19 remains unknown. This study analyzed clinical characteristics, length of stay (LOS), and in-hospital mortality of patients with HF admitted for COVID-19 at an academic tertiary care center.

## METHODS

We conducted a case control study on patients aged 18 years or older admitted to 1 of 5 Erlanger Healthcare System hospitals with COVID-19 and HF from January 2020 to February 2021. Utilizing the electronic health records, we confirmed the diagnosis of COVID-19 with positive polymerase chain reaction test and classified heart failure (HF) into heart failure with preserved ejection fraction (HFpEF), heart failure with mid-range ejection fraction (HFmrEF), or heart failure with reduced ejection fraction (HFrEF). The cardiac function was verified through echocardiogram results and supporting medical history were used for stratification according to the American Heart Association definition of HF,[13].

Further patient data collection included patient demographics, substance use, diagnoses, length of stay (LOS), number of cardiac arrests during admission, and in-hospital death. Through this manual chart review process, we identified patient factors of age, sex, body mass index (BMI), substance use, and comorbidities. We set specific comorbidities related to the cardiopulmonary system such as atrial fibrillation (AF), asthma, coronary artery disease (CAD), chronic bronchitis (CB), chronic kidney disease (CKD), chronic obstructive pulmonary disease (COPD), diabetes mellitus (DM), hyperlipidemia (HLD), hypertension (HTN), obstructive sleep apnea (OSA), peripheral arterial disease (PAD), ischemic stroke (stroke), and venous thromboembolism (VTE), and substance use. VTE included pulmonary embolism (PE) and deep venous thrombosis (DVT). Substance use included tobacco, alcohol, and intravenous drug use (IVDU).

After collecting data, we used R statistical software to determine which risk factors were associated with LOS or mortality. Risk factors (age, sex, BMI, tobacco use, alcohol use, IVDU, AF, asthma, CAD, CB, CKD, COPD, DM, HLD, HTN, OSA, PAD, stroke and VTE) were set as either categorical or continuous variables depending on the type of statistical tests applied. For categorical variables, the chi-square test was conducted. If the sample size was small, we replaced the chi-square test with the Fisher’s exact test. For continuous variables, the t-test or the Whitney’s U test was conducted. If the risk factor followed a normal distribution, a t-test was performed with mean and standard deviations. If the risk factor did not follow a normal distribution, the Whitney’s U test was performed with median and interquartile ranges (IQR).

We compared every risk factor to determine if a significant difference exists by heart failure type. In particular, age, sex, BMI, tobacco use, alcohol use, IVDU, AF, asthma, CAD, CB, CKD, COPD, DM, HLD, HTN, OSA, PAD, stroke and VTE were compared between patients with HFpEF versus patients with HFrEF. Various statistical tests were applied to compare the risk factors accordingly. The t-test was used for difference in age. The Whitney’s U test was used for differences in BMI and length of hospital stay (LOS). Chi-square test or Fisher’s exact test was used for differences in sex, substance use, comorbidities, number of cardiac arrests, and in-hospital death. Risk factors were also evaluated to determine their effects on mortality in HF patients in this study. All results with a p-value less than 0.05 were considered statistically significant.

We considered risk factors as dichotomous categorical variables when determining whether or not they were related to mortality. Risk factors, age and BMI, were converted from continuous to dichotomous categorical variables by designating specific parameters. Patients aged 65 and above (age 65+) were counted precisely, as this is the general age considered senior citizen. We defined obesity as BMI 30 and above. Including these converted categorical variables, we conducted the chi-square test to compare which risk factors were associated with in-hospital mortality. For risk factors with small sample size, we used the Fisher’s exact test instead. The corresponding odds ratio (OR), 95% confidence intervals (95% CI), and p-value were calculated for each factor. We adjusted the odds ratio (Adjusted OR) for confounding variables alcohol, tobacco, and IVDU using multivariate regression.

To maintain consistency in our study, we selected identical risk factors to test for mortality and LOS. To determine the effects of risk factors on LOS of our study population (HF patients hospitalized with COVID-19), we calculated the median length of stay for patients with and without each risk factor. Respective interquartile ranges per median were also computed. We conducted the Whitney’s U test with the resulting medians to determine whether a significant difference exists between patients with the risk factor versus patients without the risk factor. LOS was further evaluated using multivariate regression with gender and IVDU as confounding factors. P-value less than 0.05 was considered statistically significant for risk factor impact on LOS.

## RESULTS

Our study population consisted of patients with a prior diagnosis of HF admitted for COVID-19. A total of 211 patients were included in the study. Of these patients, 119 (54.5%) had HFpEF and 96 (45.5%) had HFrEF. **Table 1** outlines the clinical characteristics of our study population stratified by HF types. Compared to patients with HFrEF, patients with HFpEF were significantly more overweight or obese, more likely to have a stroke, and predominantly female. Furthermore, there was no significant difference in age, substance use, comorbidities (excluding stroke), LOS, number of cardiac arrest(s), or in-hospital death between patients with HFpEF and patients with HFrEF.

**Table 1.** Clinical characteristics and outcomes of study population according to heart failure types.

|  |  |  |  |  |
| --- | --- | --- | --- | --- |
|  | **Total** | **HFpEF**<br>**(n=115; 54.5%)** | **HFrEF**<br>**(n=96; 45.5%)** | **p-value** |
| Age, yrs | 70.4 ± 12.9 | 70.1 ± 13.7 | 70.9 ± 11.9 | 0.661 |
| Male | 119 (56.4%) | 50 (43.5%) | 69 (71.9%) | **<0.001** |
| BMI, kg/m<sup>2</sup> | 29.8 (24.8, 36.5) | 31.9 (26.0, 39.6) | 27.2 (23.5, 31.7) | **<0.001** |
| <u>Substance use</u> |  |  |  |  |
| Tobacco | 78 (37.0%) | 39 (33.9%) | 39 (40.6%) | 0.111 |
| Alcohol | 31 (14.7%) | 12 (10.4%) | 19 (19.8%) | 0.622 |
| IVDU | 14 (6.6%) | 7 (6.1%) | 7 (7.3%) | 0.084 |
| <u>Comorbidities</u> |  |  |  |  |
| AF | 100 (47.4%) | 49 (42.6%) | 51 (53.1%) | 0.166 |
| Asthma | 24 (11.4%) | 24 (11.4%) | 16 (13.9%) | 0.285 |
| CAD | 122 (57.8%) | 56 (48.7%) | 66 (68.8%) | 0.084 |
| CB | 7 (3.3%) | 4 (3.5%) | 3 (3.1%) | 0.650 |
| CKD (All stages): | 146 (69.2%) | 78 (67.8%) | 68 (70.8%) | 0.977 |
| 1 | 3 (1.4%) | 1 (0.9%) | 2 (2.1%) |  |
| 2 | 20 (9.5%) | 11 (9.6%) | 9 (9.4%) |  |
| 3 | 73 (34.6%) | 39 (33.9%) | 34 (35.4%) |  |
| 4 | 22 (10.4%) | 12 (10.4%) | 10 (10.4%) |  |
| 5 | 23 (10.9%) | 13 (11.3%) | 10 (10.4%) |  |
| COPD | 76 (36.0%) | 38 (33.0%) | 38 (39.6%) | 0.423 |
| DM | 139 (65.9%) | 80 (69.6%) | 59 (61.5%) | 0.271 |
| HLD | 117 (55.5%) | 68 (59.1%) | 49 (51.0%) | 0.301 |
| HTN | 206 (97.6%) | 112 (97.4%) | 94 (97.9%) | 0.648 |
| OSA | 48 (22.7%) | 28 (24.3%) | 20 (20.8%) | 0.544 |
| PAD | 29 (13.7%) | 11 (9.6%) | 18 (18.8%) | 0.187 |
| Stroke | 60 (28.4%) | 35 (30.4%) | 25 (26.0%) | **0.005** |
| VTE | 28 (13.3%) | 19 (16.5%) | 9 (9.4%) | 0.388 |
| LOS | 8.0 (5.0, 13.0) | 8.0 (4.0, 14.0) | 7.0 (5.0, 12.0) | 0.497 |
| <u>Cardiac arrest (n):</u> |  |  |  |  |
| 0 | 189 (89.6%) | 104 (90.4%) | 85 (88.5%) | 0.645 |
| 1 | 16 (7.6%) | 7 (6.1%) | 9 (9.4%) | 1 |
| 2 | 5 (2.4%) | 3 (2.6%) | 2 (2.1%) | 0.497 |
| 3 | 1 (0.5%) | 1 (0.9%) | 0 (0.0%) | 0.645 |
| In-hospital death | 43 (20.4%) | 23 (20.0%) | 20 (20.8%) | 1 |
| Values are n (%), mean ± standard deviation, median (interquartile range) |  |  |  |  |
| AF, atrial fibrillation; BMI, body mass index; IVDU, intravenous drug use; CAD, coronary artery disease; CB, chronic bronchitis; CKD, chronic kidney disease; COPD, chronic obstructive pulmonary disease; DM, diabetes mellitus; HLD, hyperlipidemia; HTN, hypertension; OSA, obstructive sleep apnea; PAD, peripheral arterial disease; Stroke, ischemic stroke; VTE, venous thromboembolism including deep venous thrombosis and pulmonary embolism; yrs, years. |  |  |  |  |

Risk factors were analyzed to determine any association with mortality in our study population. **Figure 1** summarizes the effect of each risk factor on in-hospital mortality. IVDU (OR: 3.33; 95% CI: 1.04 – 10.2; p = 0.035), VTE (OR: 2.53; 95% CI: 1.04 – 5.89; p = 0.035), and tobacco use (OR: 2.10; 95% CI: 1.06 – 4.15; p = 0.033) showed significant increase in risk of in-hospital death. Obesity (OR: 0.31; 95% CI: 0.15 – 0.64; p = 0.012) and OSA (OR: 0.21; 95% CI: 0.05 – 0.62; p = 0.012) showed significant decrease in risk of in-hospital death. The effect of CB on mortality remains indeterminate given 100% survival in our study (OR: 0.0; 95% CI: 1.0 – 17.5; p = 0.986). Post multivariate regression results showed age 65+ as a significant risk factor (Adjusted OR: 1.04; 95% CI: 1.00 – 1.07; p = 0.036) and increased mortality risk for VTE (Adjusted OR: 3.33; 95% CI: 1.29 – 8.43; p = 0.011). Further analysis was performed to compare the percentages of in-hospital deaths by the type of vascular disease present in our study population. **Figure 2** represents the percentages of deaths with VTE, PAD, CAD, or stroke. In our study, HF patients admitted for COVID-19 with diagnosis of VTE, PAD, CAD, or stroke faced in hospital death by 36%, 31%, 24%, and 18%, respectively.

**Figure 1.**
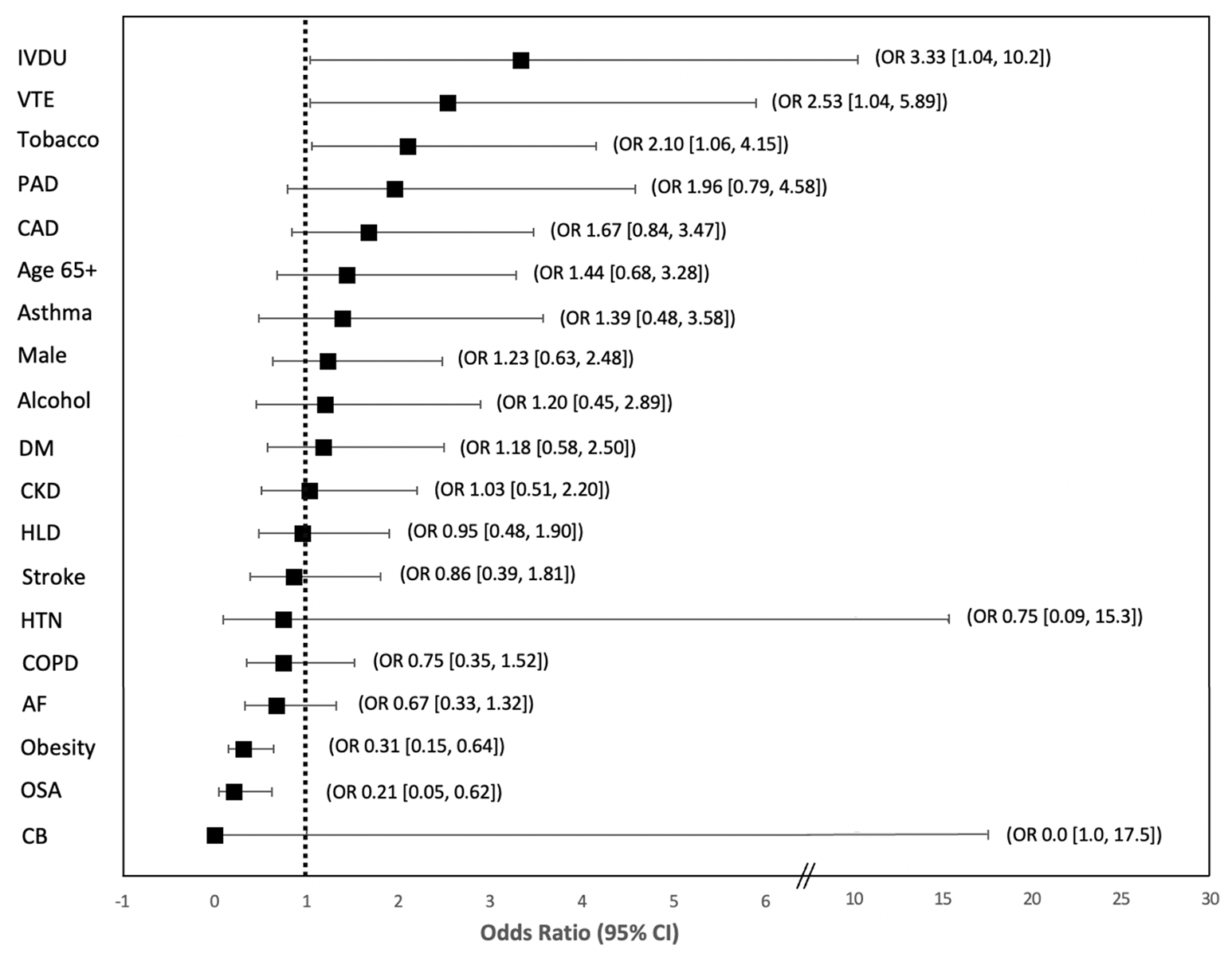
Forest plot for the effect of risk factors on in-hospital mortality for all patients with HF admitted for COVID-19. OR, odds ratio; CI, confidence interval; IVDU, intravenous drug use; VTE, deep venous thrombosis and pulmonary embolism; PAD, peripheral arterial disease; CAD, coronary artery disease; Age 65+, age greater than or equal to 65; DM, diabetes mellitus; CKD, chronic kidney disease; HLD, hyperlipidemia; HTN, hypertension; COPD, chronic pulmonary obstructive disease; AF, atrial fibrillation; OSA, obstructive sleep apnea; CB, chronic bronchitis;

**Figure 2.**
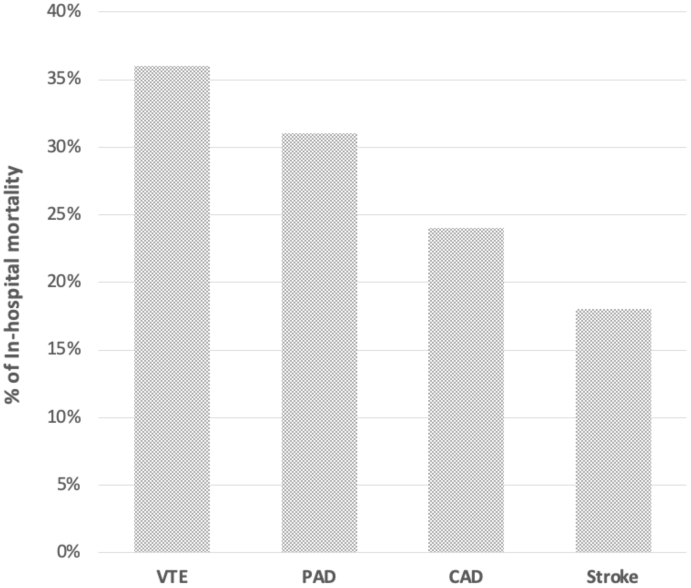
In-hospital mortality of HF patients stratified by type of vascular disease

Risk factor association with LOS was also compared in our study. **Table 2** outlines the effect of each risk factors on hospital LOS. IVDU, asthma, stroke, and PAD significantly increased LOS in our study population. The median LOS for our study population with and without IVDU was 14 days (IQR: 11 to 19 days) and 7 days (IQR: 4.5 to 12 days), respectively. The median LOS for patients with and without asthma was 13 days (IQR: 9 to 21 days) and 7 days (IQR 4 to 12 days), respectively. The median LOS for patients with and without stroke was 11 days (6 to 18 days) and 7 days (4 to 11 days), respectively. The median LOS for patients with and without PAD was 9 days (IQR: 6 to 16 days) and 7 days (IQR: 5 to 12 days), respectively. Interestingly, females had significantly longer LOS than males. The median LOS for females and males was 9 days (IQR: 5 to 14 days) and 7 days (IQR: 4 to 12 days), respectively.

**Table 2.** Length of hospital stay for patients with HF admitted for COVID-19 by risk factors.

|  |  |  |  |
| --- | --- | --- | --- |
| **Risk factors** | **LOS (+)** | **LOS (-)** | **p-value** |
| Age ≥ 65 | 8 (5, 12) | 8 (5, 14) | 0.439 |
| Male | 7 (4, 12) | 9 (5, 14) | **0.036** |
| Obesity (BMI ≥ 30) | 8 (5, 12) | 8 (5, 13) | 0.810 |
| <u>Substance use:</u> |  |  |  |
| Tobacco | 7 (4, 13) | 8 (5, 13) | 0.507 |
| Alcohol | 10 (5.5, 14.5) | 8 (5, 12) | 0.260 |
| IVDU | 14 (11.3, 19.3) | 7 (4.5, 12) | **0.002** |
| <u>Comorbidities:</u> |  |  |  |
| AF | 8 (5, 12) | 8 (4.5, 14) | 0.859 |
| Asthma | 13 (9, 20.8) | 7 (4, 12) | **0.001** |
| CB | 5 (5, 11) | 8 (5, 13) | 0.472 |
| CAD | 8 (5, 13) | 8 (4, 13) | 0.817 |
| CKD | 8 (4, 12) | 7 (5, 14) | 0.716 |
| COPD | 7 (5, 11) | 8 (5, 14) | 0.247 |
| DM | 9 (4.5, 14) | 7 (5, 11.5) | 0.428 |
| HLD | 7 (5, 12) | 9 (5, 14) | 0.203 |
| HTN | 8 (5, 13) | 12.5 (4, 20.5) | 0.901 |
| OSA | 10 (5, 14) | 7.5 (5, 13) | 0.501 |
| PAD | 9 (6, 16) | 7 (5, 12) | **0.01\*** |
| Stroke | 11 (5.75, 17.5) | 7 (4, 11) | **0.002** |
| VTE | 11.5 (5, 14.3) | 7 (5, 12) | 0.127 |
| Values are represented by median (interquartile range) number of days hospitalized. |  |  |  |
| AF, atrial fibrillation; BMI, body mass index (kg/m<sup>2</sup>); CAD, coronary artery disease; CB, chronic bronchitis; CKD, chronic kidney disease; COPD, chronic obstructive pulmonary disease; DM, diabetes mellitus; HLD, hyperlipidemia; IVDU, intravenous drug use; Stroke, ischemic stroke; VTE, venous thromboembolism including deep venous thrombosis and pulmonary embolism. |  |  |  |
| *: adjusted for gender and IVDU |  |  |  |

## DISCUSSION

HF is a condition that reduces immunity. Through the increase of tumor necrosis factor alpha (TNF-a) and decrease of interleukin-10, HF promotes an inflammatory process that intensifies the severity of infectious illness,[2]. COVID-19 was not an exception to the infections that exacerbate patients with HF. Our study revealed that 20% of patients with prior HF hospitalized with COVID-19 faced in-hospital death. Compared to a previous study that tallied in-hospital deaths in 1000 hospitals, this percentage was vastly greater than 4.5% of patients admitted without COVID-19 or HF and consistent with 24% of the patients hospitalized with COVID-19 with prior HF,[3]. Our study was also congruent for no significant difference in mortality between HF types with a previous study that indicated twice the risk of mortality in COVID-19 patients with HF versus without HF,[1]. We designated comorbidities AF, HTN, DM, CKD, COPD, OSA, CB, asthma, HLD, stroke, CAD, PAD, and VTE to investigate their effects on LOS and mortality of HF patients admitted with COVID-19. Through this process, we discovered that the diagnosis of obesity resulted in 3 times less risk of death than without obesity (OR 0.31; 95% CI, 0.15 – 0.64; p = 0.002). This finding was consistent with a meta-analysis study that revealed being overweight and obese were protective factors against cardiovascular mortality and all-cause mortality in patients with HF,[8]. We also found that VTE resulted in greater than three times the risk of death in COVID-19 patients with HF than without VTE (OR 3.33; 95% CI, 1.29 – 8.43; p = 0.011). More specifically, our study revealed that 1 of 3 patients with HF, VTE, and COVID-19 had in-hospital death while previous research showed that 1 of 5 patients with HF, VTE, and no COVID-19 had in-hospital death,[10].

The mechanism of how COVID-19 increases mortality in patients with HF and VTE remains unknown. However, understanding the pathophysiology of HF and COVID-19 helps develop a conjecture about the increased mortality in patients with HF, COVID-19, and VTE as reflected in our study. HF promotes thrombogenesis through endothelial dysfunction and inflammation, as noted by decreased endothelial nitric oxide synthase activity and increased interleukin-1, interleukin-6, TNF-a, soluble thrombomodulin, circulating endothelial cells, C-reactive protein, angiopoietin-2, tissue factor, E-selectin, and von Willebrand factor (vWF),[5]. Moreover, HF reduces hemodynamic performance, exacerbating the ability to cope with increased cardiac output required in the setting of inflammation due to infections,[2]. COVID-19 induced acute respiratory distress syndrome (ARDS) showed significantly higher platelets, fibrinogen, and thrombotic complications than non-COVID-19 ARDS,[4]. Furthermore, COVID-19 ARDS patients had more than 5 times incidence of PE with very high levels of vWF antigen,[4].

Perhaps, the increased vWF in both HF and COVID-19 provides an explanation to the increased mortality of patients with COVID-19, HF, and VTE versus patients with COVID-19 and VTE, HF and VTE, or COVID-19 without VTE. The percentage of death for patients with COVID-19, HF, and VTE in our study was 35.7% while the percentage of death for patients with COVID-19 and VTE, HF and VTE, and COVID-19 without VTE was approximately 25%, 20% and 13% respectively in other studies,[3,7,10]. Increased mortality of patients inclusive of all three conditions, VTE, HF, and COVID-19, compared to patients with an exclusion of any of these conditions demonstrates that thrombotic parameters from combination of HF and COVID-19 warrants investigation. Supplemental to these findings, studies showed that patients with COVID-19 and HF had twice the greater risk of death compared to patients with COVID-19 and no HF while patients with COVID-19 and VTE also had twice the greater risk of death compared to patients with COVID-19 and no VTE,[1,7]. Studies also revealed that VTE increased the incidence of HF while HF also increased the incidence of VTE, highlighting the detrimental cycle of HF in association with VTE,[5,9]. These relations of COVID-19, HF, and VTE underscore the importance of optimal medical management for morbidity and mortality reduction.

The exact treatment regimen for acute COVID-19 infection is yet to be established. The treatment decisions become even more abstruse in the setting of a coexistent heart failure. All patients in this study had prior HF and received anticoagulation. Patients with deep venous thromboembolism, pulmonary embolus, or AF with CHA2DS2-VASc score greater than or equal to 2 received various types of anticoagulation at treatment dose while patients without these conditions received heparin or enoxaparin at thromboprophylaxis dose. All patients in this study with CAD, PAD, and ischemic stroke received statin and antiplatelet therapy with aspirin or other P2Y12 inhibitors. Our study also showed HF patients admitted for COVID-19 had longer LOS with diagnosis of PAD or ischemic stroke (7 days vs. 9 days; p = 0.012 and 7 days vs. 11 days, p < 0.001; respectively). A multihospital retrospective study revealed that antiplatelet and statin use decreased VTE or mortality by 35% and 45%, respectively in COVID-19 patients,[11]. Another study also showed that COVID-19 patients receiving therapeutic anticoagulation prior to admission had three times less risk of developing VTEs and did not significantly suffer more from all-cause death than patients without anticoagulation^7^. Furthermore, heparin use during admission was associated with twice as less mortality in HF patients with COVID-19,[12].

### Limitations

The use of electronic health records with ICD-9/10 codes for HF potentially misclassified some patients per HF type. ICD-9/10 codes with congestive heart failure, concurrent HFpEF and HFrEF, and HF without further specifications required sorting to appropriate HF types. To minimize these classification errors, we manually ascertained HF type through comprehensive review of documentations, echocardiograms, and medication list. HFmrEF was not analyzed as a separate entity due to inadequate number. In efforts to ensure the absence of HFmrEF category does not derange our study, we performed analyses with HFmrEF classified as HFpEF and without HFmrEF. The significance of the results did not vary when comparing HFmrEF as HFpEF or excluding the HFmrEF patient data sets altogether. Interestingly, our study showed OSA to be a protective factor against mortality (OR 0.21; 95% CI, 0.05 – 0.62; p = 0.012). However, the significance of OSA for mortality reduction remains questionable in our study given its low prevalence in our study population. We identified risk factors for OSA including BMI, HTN, smoking, asthma, sex (male), and DM. HTN, DM, sex (male), obesity, tobacco use, and asthma were present in 98%, 66%, 56%, 49%, 36%, and 11% of our patients, respectively. Given the predominant existence of OSA risk factors and only 3 OSA patients with in-hospital death, we predict that OSA is under diagnosed in our study population. Future studies should also explore why female patients with HF hospitalized with COVID-19 had significantly higher LOS than male patients with HF hospitalized with COVID-19.

## CONCLUSION

Our study revealed that patients with prior HF and VTE admitted for acute COVID-19 infection faced greater than 3-fold increased risk of in-hospital death versus patients without VTE. Our study also showed that increased hospital LOS in patients with stroke and peripheral arterial disease. Standardized antithrombotic treatment for patients with HF and COVID-19 are inconclusive. With several studies demonstrating mortality decrease with antiplatelet, anticoagulation, and statin therapy for HF patients hospitalized with COVID-19, clinicians could possibly consider a lower threshold in initiating antithrombotic and statin therapy if findings are confirmed in other populations.

## Data Availability

Data are available within the article or its supplementary materials

## Notes

### Competing Interest Statement

The authors have declared no competing interest.

### Funding Statement

The authors received no financial support for the research, authorship, and/or publication of this article.

### Author Declarations

University of Tennessee at Chattanooga Institutional Review Board

